# Age-adjusted mortality from systolic heart failure (HFrEF) is almost double that of diastolic heart failure (HFpEF) over recent years, with significant racial disparity

**DOI:** 10.1101/2025.10.08.25337632

**Authors:** Mohammad Reza Movahed, Harrison Feerst, Mehrtash Hashemzadeh

## Abstract

**Background:** Heart failure remains a leading cause of morbidity and mortality in the United States. Systolic heart failure (SHF; HFrEF) used to be considered as more lethal than diastolic heart failure (DHF; HFpEF). This study evaluates national age-adjusted mortality trends for SHF and DHF from 2016 to 2020, with particular attention to sex- and race-based differences.

**Methods:** We performed a retrospective analysis of adults ≥20 years hospitalized with SHF or DHF in the Healthcare Cost and Utilization Project National Inpatient Sample from 2016 to 2020. Age-adjusted mortality rates per 100,000 population were calculated using direct standardization to the 2000 U.S. standard population. Trends were stratified by sex and race to identify demographic disparities.

**Results:** We identified 7.36 million SHF and 10.06 million DHF hospitalizations between 2016 and 2020. SHF maintained higher absolute mortality throughout the study, almost double that of diastolic heart failure (250.4 to 328.6 per 100,000; +31.2%), but DHF mortality rose more sharply in relative terms (164.1 to 225.8 per 100,000; +37.6%). From 2016–2019, mortality rates were relatively stable, but between 2019 and 2020, SHF mortality increased by 27.8% and DHF mortality by 33.5%. Minority populations experienced the steepest mortality surges, particularly Native American, Hispanic, and Black patients, narrowing their historical mortality gaps between SHF and DHF. Similarly, male DHF mortality increased disproportionately in 2020 (47.7% rise vs. 22.2% for females).

**Conclusions:** DHF does represent the predominant heart failure phenotype in the United States in terms of hospitalization occurrences, and shows a faster growth trajectory in mortality relative to SHF. Importantly, however, SHF remains more lethal in absolute terms and has not come close to being surpassed by DHF despite a sharp rise in DHF mortality during 2020.

## Introduction

Heart failure (HF) is a progressive clinical syndrome with a high morbidity and mortality, affecting over 6.7 million adults in the United States [1]. Although the incidence of disease has been showing signs of stabilization in more industrialized countries such as the United States, prevalence has continued to increase alongside an aging population [2-4]. Historically, heart failure has been categorized based on left ventricular ejection fraction (LVEF), with systolic heart failure (SHF), also known as heart failure with reduced ejection fraction (HFrEF), having a LVEF ≤40%, and diastolic heart failure (DHF), also referred to as heart failure with preserved ejection fraction (HFpEF), having a LVEF >50% [5-8]. Although there is also a recognized intermediate classification of HF with mid-range ejection fraction (HFmrEF), defined as a LVEF between 40% and 50%, for the purposes of this study we will focus only on the two previously mentioned main categories, SHF and DHF [6, 7].

Systolic heart failure is defined by impaired ventricular contraction, resulting in a reduced ability of the left ventricle to eject blood during systole, and a subsequent increased end-diastolic volume [5]. Dilation allows for the ventricle to accommodate the increased volume initially, but over time, cardiac remodeling and progressive eccentric hypertrophy eventually exceed the heart’s ability to compensate, resulting in heart failure [5, 8]. The underlying pathophysiology typically involves an initial myocardial insult such as ischemic injury, pressure overload, or cardiomyopathy, all of which impair contractile force and reduce cardiac output. The body reacts to this reduced output by activating the renin-angiotensin-aldosterone system (RAAS) in the kidneys to increase preload, as well as the sympathetic nervous system to intensify contractility, which results in a cycle of dysfunction that causes the progressive dilation that is seen [9]. Clinically, this presents signs of fluid congestion and retention, such as dyspnea and peripheral edema, as well as fatigue and exercise intolerance [10].

Diastolic heart failure has many of the same symptoms as SHF but differs significantly in the primary cause and physiological changes seen. In DHF, the heart has impaired ventricular relaxation and is unable to accommodate an appropriate amount of blood volume. Concentric hypertrophy in the myocardium reduces the space in the cardiac chamber and prevents significant dilation from occurring, which in turn reduces the stroke volume [11, 12]. A reduced stroke volume causes the heart to compensate by increasing its contraction rate, and over time, that increased work and stress exacerbate the hypertrophy and worsen the heart failure [11, 12]. Clinically, this presents with exertional dyspnea, peripheral edema, and fatigue, and is most easily distinguished from systolic heart failure through evaluation of the LVEF [4]. The pathology behind DHF is associated with stress over time, with a multisystem process of cardiac age, hypertension, diabetes, obesity, and other cardiometabolic disorders contributing to inflammation, fibrosis, and stiffness of the heart, all leading to DHF [13].

Current evidence suggests that systolic heart failure and diastolic heart failure have broadly similar long-term outcomes, with major differences being mainly in the underlying causes of death rather than in overall rates [1, 14]. The primary distinction lies not in the total mortality burden but in its cause: SHF is linked more often to direct cardiovascular deaths, whereas DHF patients die more frequently of non-cardiovascular conditions related to advanced age and comorbidities [1, 14]. Although mortality and morbidity have improved modestly for both phenotypes over time, SHF has benefited more from the advent of disease-modifying therapies, while DHF outcomes remain constrained by the absence of proven mortality-reducing treatments and the high prevalence of non-cardiac comorbidities [13, 15, 16].

Sex and race also exert important influences on the epidemiology and outcomes of heart failure phenotypes. Numerous studies have documented that men are disproportionately affected by systolic heart failure, whereas women are more likely to develop diastolic heart failure [4, 17]. The prevailing explanation is that men have a higher lifetime prevalence of coronary artery disease and myocardial infarction: conditions that directly damage myocardial contractility and predispose to SHF. In contrast, women more frequently have risk factors such as hypertension, obesity, and metabolic syndrome that promote concentric left ventricular remodeling and impaired diastolic relaxation [4, 17]. Mortality rates between genders are similar in DHF, but women tend to have a better prognostic outlook in SHF [18]. Marked racial differences have also been made apparent in literature. Analyses of large national datasets show that Black individuals have the highest age-adjusted mortality rates for both DHF and SHF in the United States, followed by White individuals, whereas Hispanic and Asian populations have the lowest rates [1, 19]. These disparities likely reflect a combination of factors, including a higher prevalence of hypertension and cardiometabolic disease, differences in access to preventive and specialty care, socioeconomic stressors, and structural inequities that can influence both incidence and outcomes.

In this study, we aimed to analyze national data on the incidence and age-adjusted mortality of SHF vs DHF from 2016 to 2020, with particular emphasis on changes over time and demographic subgroups.

## Methods

### Statistical Analysis

All analyses were conducted using data from adults (≥20 years) hospitalized between 2016 to 2020. A retrospective analysis was conducted using data from the Healthcare Cost and Utilization Project (HCUP) National Inpatient Sample (NIS) from 2016 to 2020. The NIS is the largest publicly available all-payer inpatient healthcare database in the United States and approximates a 20% stratified sample of all U.S. community hospital discharges, allowing for weighted national estimates.

Annual frequencies of SHF and DHF were identified using International Classification of Diseases, Tenth Revision (ICD-10) diagnosis codes. To account for changes in population structure over time, age-adjusted rates per 100,000 population were computed using the direct standardization method, with the 2000 U.S. standard population set as the reference. Age adjustment was performed using standard age groupings consistent with CDC guidelines. Data was analyzed using STATA 19 (Stata Corporation, College Station, TX), incorporating NIS sampling design elements, including discharge weight, to generate nationally representative estimates.

## Results

A total of 7,364,023 patients with systolic heart failure (SHF) and 10,064,223 patients with diastolic heart failure (DHF) between the years 2016 and 2020 were included in the dataset analyzed for this study. The average age of patients with SHF and DHF was 69.25±14.12 and 73.64±12.78, respectively. Males comprised the majority, 63.76%, of SHF, and females comprised the majority, 58.98%, of DHF. Table 1 summarizes the demographic characteristics of the patient population analyzed for this study.

**Table 1:** Base Data and Demographic Information.

|  | SHF | DHF | SHF (Mortality) | DHF (Mortality) |
| --- | --- | --- | --- | --- |
| Average Hospitalization Frequency | 294,555 | 402,561 | 15,727 | 16,743 |
| Mean Age ± SD | 69.25±14.12 | 73.64±12.78 | 73.10±13.02 | 76.93±11.46 |
| Median Age (IQR) | 71(60-80) | 75(66-84) | 75(65-83) | 79(70-87) |
| Gender |  |  |  |  |
| Male | 63.76% | 41.02% | 63.75% | 42.62% |
| Female | 36.24% | 58.98% | 36.25% | 57.38% |
| Race |  |  |  |  |
| White | 66.49% | 72.63% | 70.47% | 74.77% |
| Black | 19.85% | 15.80% | 15.35% | 13.13% |
| Hispanic | 8.33% | 6.98% | 7.92% | 6.71% |
| Asian/Pac Isl | 2.13% | 2.03% | 2.65% | 2.57% |
| Native American | 0.59% | 0.48% | 0.66% | 0.52% |
| Others | 2.61% | 2.08% | 2.95% | 2.30% |

From 2016 to 2020, age-adjusted SHF remained more lethal in absolute terms throughout all five years, by a significant margin. Overall, SHF mortality exceeded DHF mortality by about 95.2 deaths per 100,000, with the absolute difference in mortality remaining steady (85–105 per 100,000) across the five years. Age-adjusted SHF mortality increased from 250.4 to 328.6 per 100,000 between 2016 and 2020, a 31.2% increase. Over the same period, DHF mortality rose from 164.1 to 225.8 per 100,000, marking a 37.6% relative increase. However, much of the mortality increase came solely from the 2020 data points. When analyzing only from 2016 to 2019, SHF mortality increased from 250.4 per 100,000 in 2016 to 257.1 in 2019, a 2.7% rise over the four years, while DHF mortality rose from 164.1 to 169.1 per 100,000, reflecting a 3.0% increase. From 2019 to 2020 alone, there was a 27.8% increase in SHF mortality and a 33.5% increase in DHF mortality (Figure 1).

**Figure 1:**
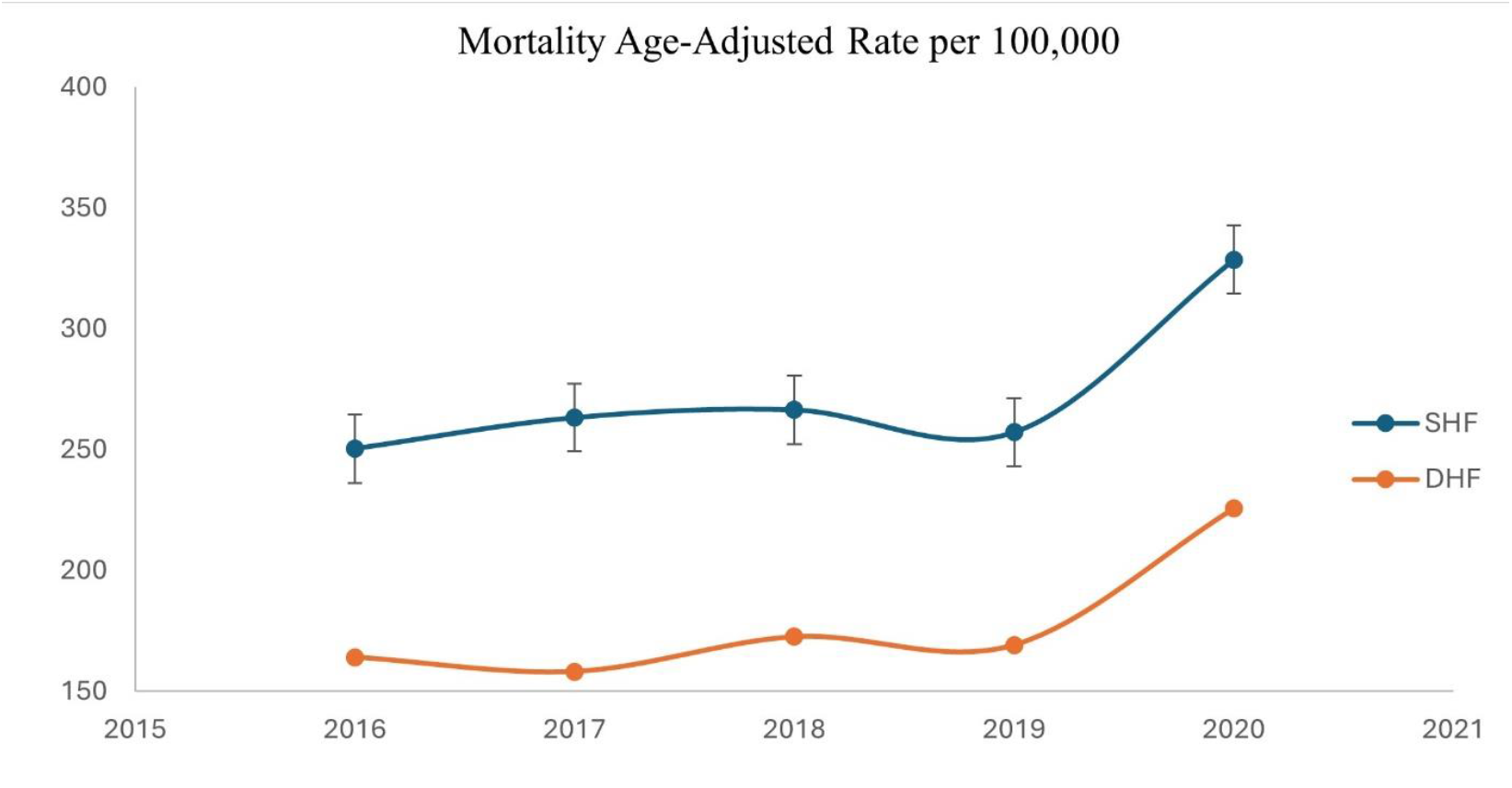
Year-on-year comparison of mortality of SHF and DHF, age-adjusted.

When stratified by gender, diastolic heart failure mortality varies modestly between genders, with no consistent predominance. Female DHF mortality increased from 156.7 per 100,000 in 2016 to 203.4 per 100,000 in 2020. Male DHF mortality rose from 172.9 to 255.9 per 100,000 during the same period. However, from 2019 to 2020, a sharp divergence occurred: male DHF mortality rose by 47.7%, compared to only a 22.2% increase in females, resulting in the highest male DHF mortality observed across the study period. SHF mortality increased for both sexes as well, with female mortality rising from 254.8 to 326.6 per 100,000, and male mortality rising from 250.1 to 334.5 per 100,000 over the 5 years, with no significant disparity between the sexes at any year. However, once again, a significant increase in mortality is seen in 2020, from 255.9 to 334.5 per 100,000, a 30.7% increase, seen in males, and from 263.7 to 326.6 per 100,000, a 23.8% increase, seen in females (Figure 2).

**Figure 2:**
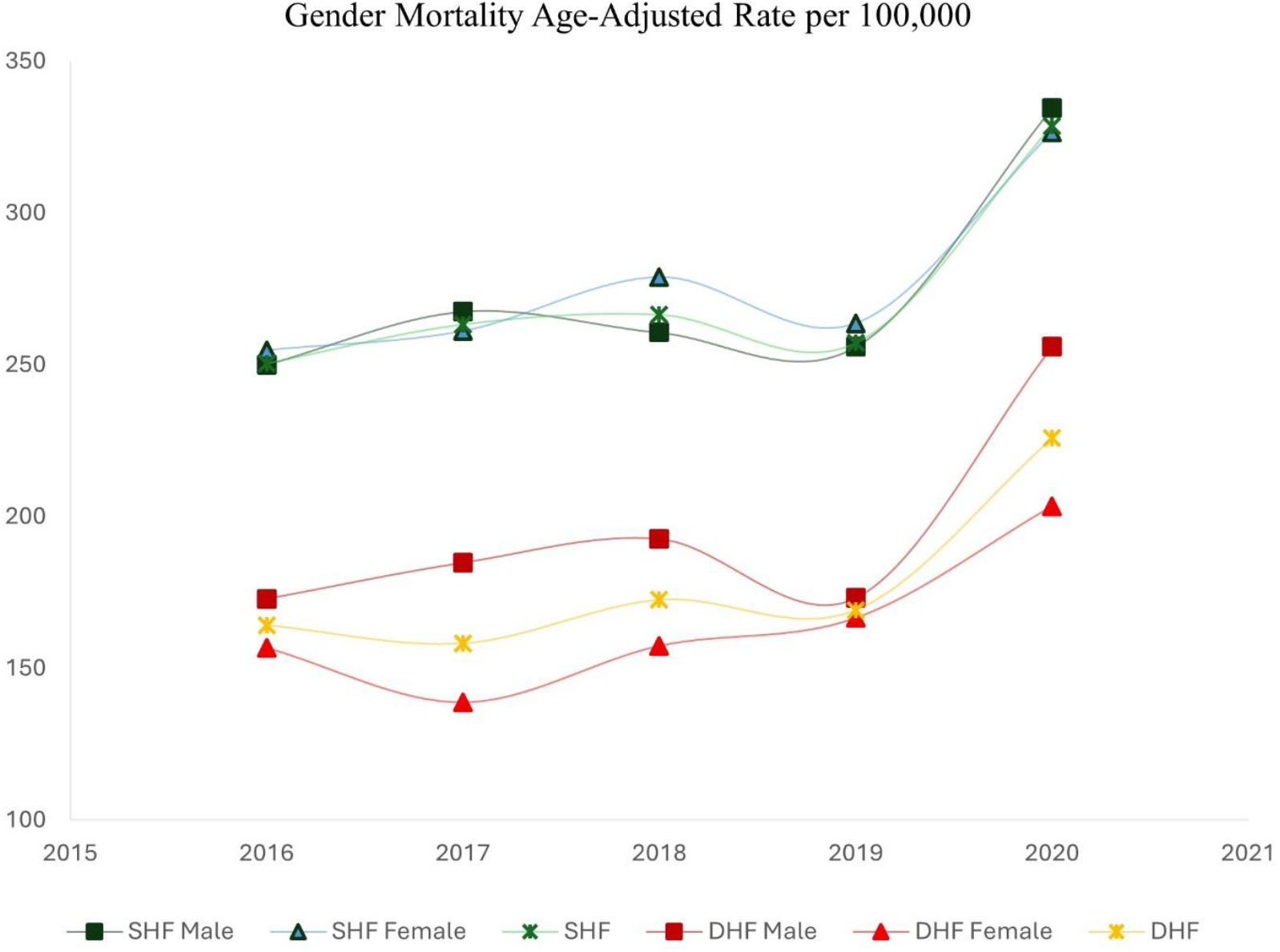
Year-on-year comparison of mortality of SHF and DHF stratified for gender, age-adjusted.

Mortality exhibited significant racial disparities and disproportionate increases, particularly between 2019 and 2020. Although SHF mortality remained consistently higher across all racial groups, DHF mortality increased more rapidly in several racial populations, particularly in Black, Hispanic, Native American, and other groups, narrowing the mortality gap that previously existed between the two heart failure subtypes. Native American patients showed the highest overall rise in DHF mortality, from 133.8 to 335.1, a 150.4% increase, alongside a 77.2% increase in SHF mortality, 289.4 to 512.6. Hispanic individuals also experienced a drastic 82.6% increase in DHF mortality, 155.3 to 283.6, and a 40.9% increase in SHF mortality, 249.9 to 352.1. In White individuals, DHF mortality increased from 188.1 to 226.9, a 20.6% rise, while SHF mortality rose from 272.0 to 354.6 per 100,000, a 30.4% increase. In Black individuals, DHF mortality rose more sharply, from 131.1 to 208.0, representing a 58.6% increase, with SHF mortality increasing similarly from 212.9 to 279.5, a 31.3% change. In Asian populations, DHF mortality reversed its trend, declining slightly by 2.8%, from 242.4 to 235.6, while SHF mortality rose from 304.5 to 412.7 for a 35.6% increase. For individuals categorized as Other, DHF mortality jumped 105.8% from 140.1 to 288.3, and SHF mortality rose 53.6% from 271.0 to 416.1 (Figures 3-5).

**Figure 3:**
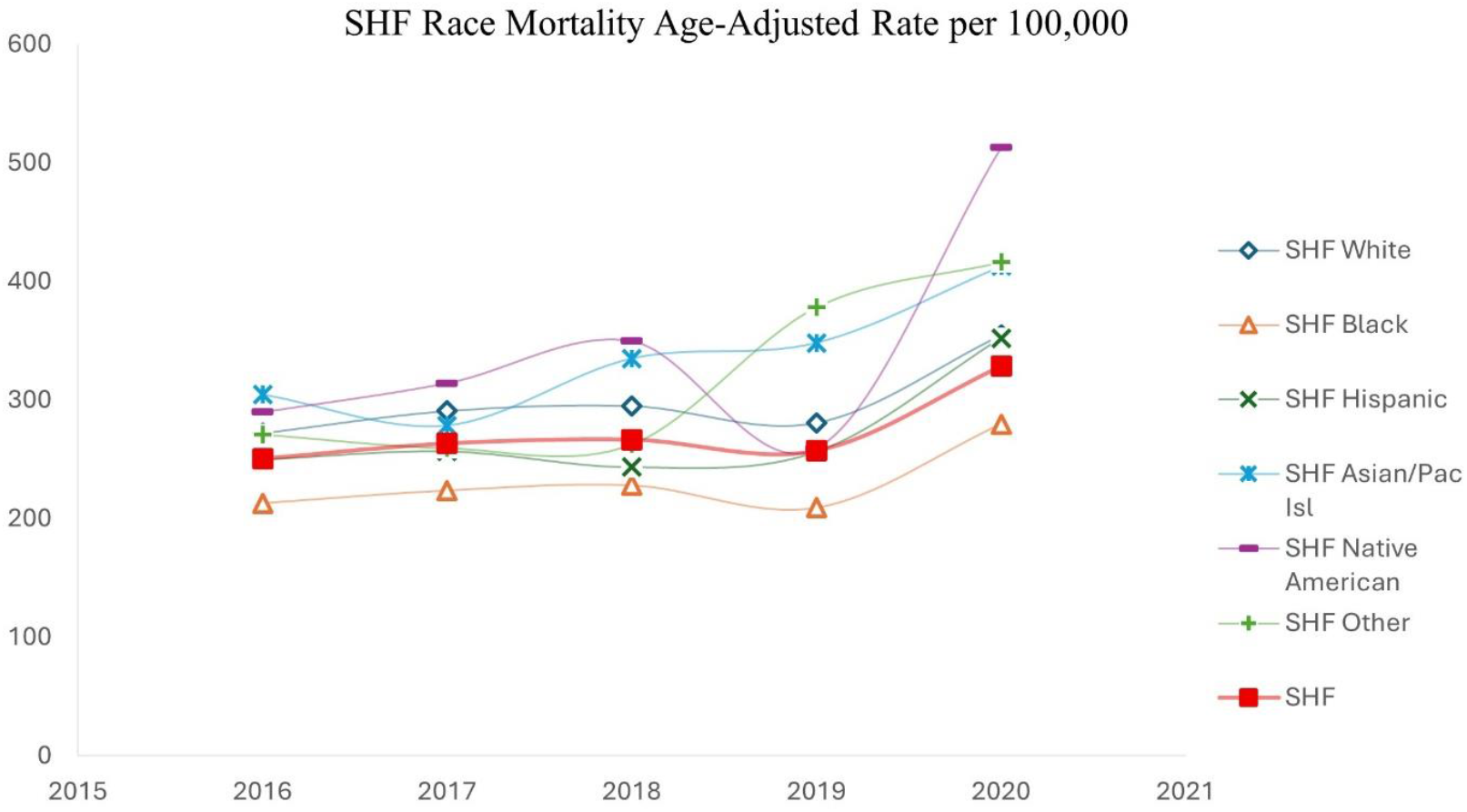
Year-on-year comparison of mortality of SHF stratified for race, age-adjusted.

**Figure 4:**
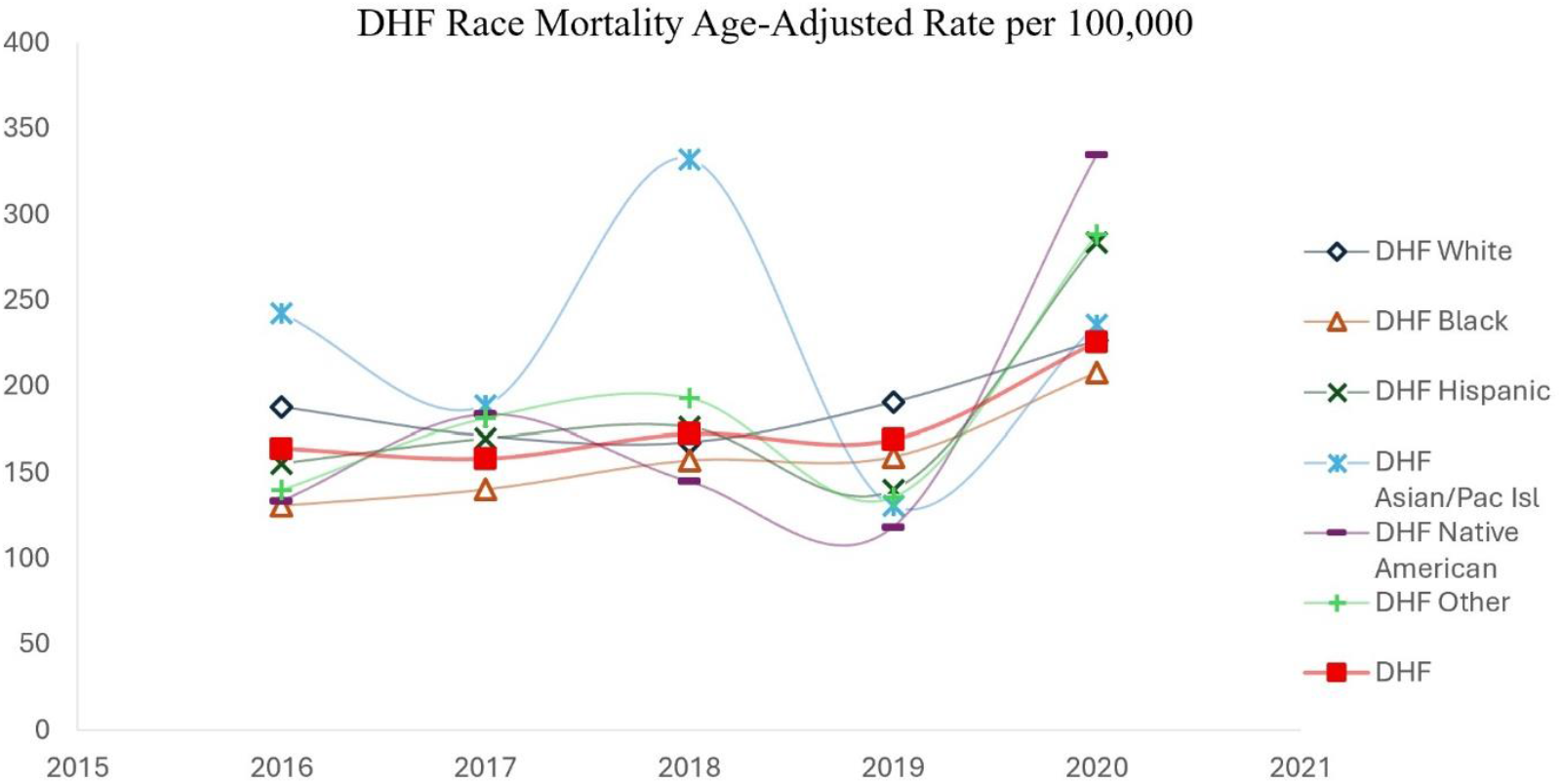
Year-on-year comparison of mortality of DHF stratified for race, age-adjusted.

**Figure 5:**
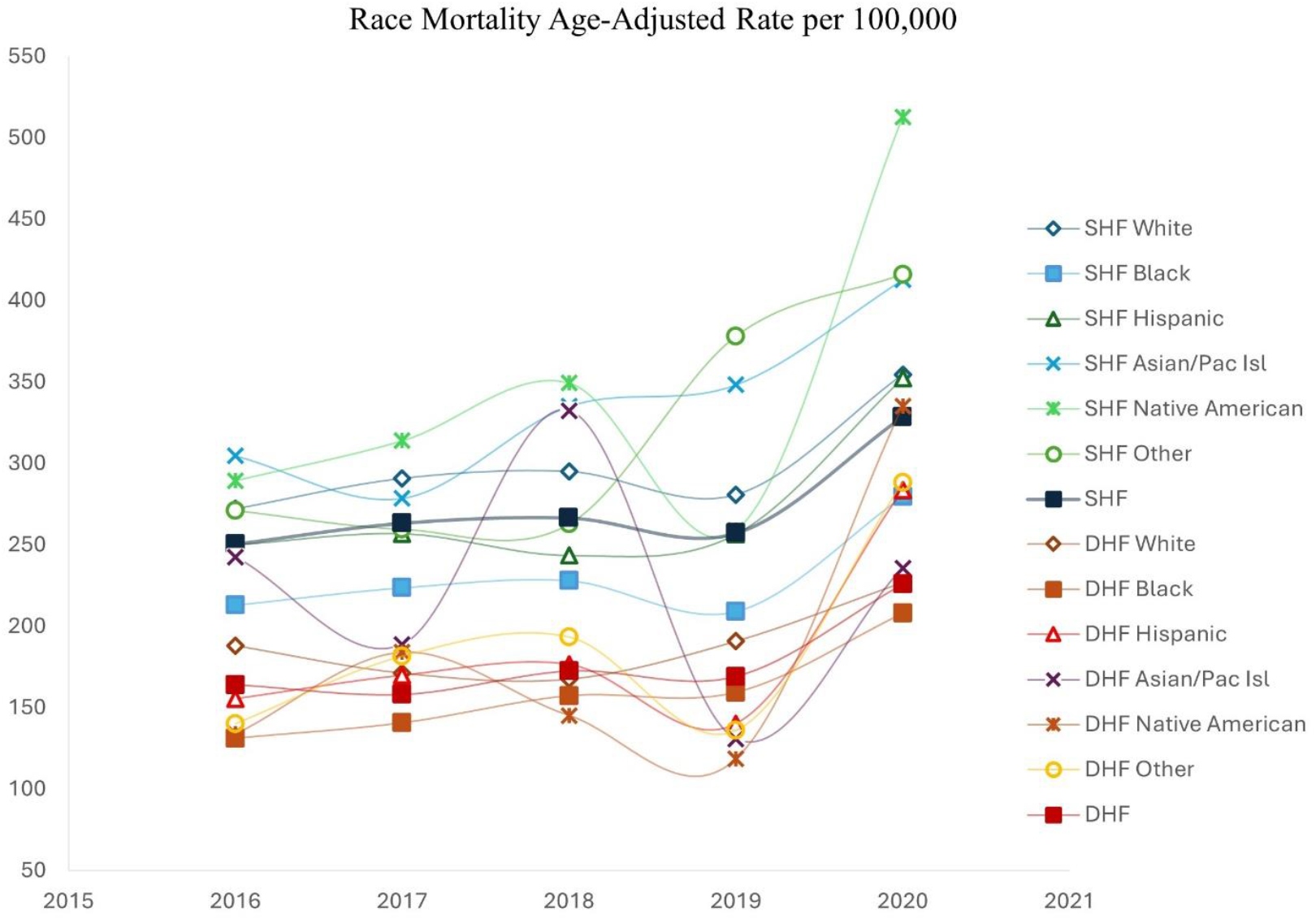
Year-on-year comparison of mortality of SHF and DHF stratified for race, age-adjusted.

## Discussion

This study evaluated mortality trends in diastolic and systolic heart failure from 2016 to 2020, with particular focus on the effects of sex and race. A key strength of our analysis was the use of age-adjusted mortality rates to compare systolic heart failure (SHF) and diastolic heart failure (DHF). Because aging is strongly linked to the mortality of heart failure, age adjustment is essential for producing valid comparisons across populations with differing demographic profiles. Without age adjustment, observed differences in mortality between racial or sex-based groups could be driven largely by differences in age structure rather than true variation in disease burden, resulting in spurious associations. By standardizing age, our analysis allowed a clearer assessment of temporal trends and demonstrated that there remains a drastic difference in mortality between SHF and DHF. Additionally, age-adjustment allows us to observe that the striking increases in SHF and DHF mortality observed in 2020 were not simply the consequence of an aging population but reflected a real and disproportionate rise in deaths. This is also particularly relevant in the context of external stressors such as the COVID-19 pandemic, in which excess mortality might otherwise be misattributed to demographic shifts. Age-adjusted mortality metrics are therefore indispensable for fair, interpretable comparisons in epidemiologic studies and strengthen the ability of this study to demonstrate meaningful changes in heart failure outcomes over time.

Most contemporary studies and reviews have concluded that systolic heart failure and diastolic heart failure have broadly similar long-term mortality, with differences thought to lie more in the causes of death than in absolute mortality rates [1, 14, 20]. Large registry analyses and meta-analyses consistently report near-identical one- and five-year survival rates between SHF and DHF, suggesting that the once-widely reported survival gap has narrowed over time and that DHF may now rival SHF in lethality [1, 20]. These findings have shaped a prevailing perception that, while the two phenotypes differ in pathophysiology, their overall mortality burdens have converged in the modern era, particularly as evidence-based therapies for SHF have matured and comorbidity-driven non-cardiac deaths in DHF have risen [13, 15, 16]. However, most of these studies rely on crude mortality counts or inadequately age-adjusted estimates, which can obscure true differences because DHF disproportionately affects older adults with inherently higher baseline mortality risk. Without standardizing for age, DHF may appear to be as lethal as SHF simply due to demographic differences rather than actual differences in disease-related mortality. In contrast, our nationally representative, age-adjusted analysis demonstrates a markedly different picture. Across more than 17 million combined SHF and DHF hospitalizations from 2016 to 2020, SHF consistently maintained a substantially higher mortality rate than DHF, with SHF showing 1.5 times more lethality in absolute terms in every year studied. Even after controlling for factors that may inflate crude death counts, such as the older age and heavier comorbidity burden of DHF patients, SHF mortality exceeded DHF mortality by an average of 95 deaths per 100,000 population throughout the five years. While DHF mortality rose more steeply in relative terms, particularly in 2020, its absolute risk of death never approached that of SHF. These findings directly challenge the prevailing narrative of mortality equivalence between SHF and DHF, underscoring the importance of rigorous age adjustment and population-level analyses when comparing heart failure phenotypes. By revealing that systolic dysfunction continues to carry a uniquely high risk of death, our data calls for a reassessment of how mortality risks in SHF and DHF are evaluated and understood

It is also important to note that nearly all of the increase in mortality seen occurred in the final year of the dataset. From 2016 to 2019, mortality rates for both phenotypes remained relatively stable, with SHF mortality rising by 2.7% and DHF mortality by 3.0%. The abrupt rise from 2019 to 2020, 27.8% for SHF and 33.5% for DHF, strongly suggests that an exogenous influence, most plausibly the COVID-19 pandemic, played a key role in driving excess mortality during 2020, likely through disruptions in chronic disease management, reduced access to care, and higher vulnerability to severe infection in patients with existing cardiovascular disease. While causation cannot be determined from this data alone, the timing meaningfully coincides with the onset of the COVID-19 pandemic, and patients with heart failure are known to be at high risk for severe COVID-19 outcomes [21]. Age adjustment fundamentally altered the interpretation of mortality trends between systolic heart failure and diastolic heart failure. On initial review of crude mortality counts, DHF appeared to account for a greater overall burden, with a higher absolute number of deaths than SHF throughout the study period. However, the age distribution of affected populations heavily influences the results seen, as DHF disproportionately affects older adults, a demographic with inherently higher baseline mortality risk, whereas SHF occurs more frequently across a broader age range, including younger individuals.

Additionally, this study reveals nuanced gender-specific differences in mortality trends between systolic heart failure (SHF) and diastolic heart failure (DHF) over the 2016–2020 period. While overall DHF mortality increased for both males and females, the pattern of change and its magnitude differed, particularly in 2020. Over the full five-year period, DHF mortality in males rose from 172.9 to 255.9 per 100,000, a 48.0% increase, compared to a 29.8% increase in females, from 156.7 to 203.4 per 100,000. However, a striking gender divergence emerged in the final year: from 2019 to 2020 alone, male DHF mortality increased by 47.7%, while female mortality rose by 22.2%. This shift resulted in the highest male DHF mortality rate of the study period, exceeding that of females by a substantial margin. Before 2020, DHF mortality rates had remained relatively close between genders, with no consistent predominance. In contrast, SHF mortality increased steadily in both sexes over the study period, with a 28.2% rise in females and a 33.7% rise in males. Importantly, SHF mortality remained slightly higher in males throughout, but the difference between sexes was modest and consistent, even during 2020 when male mortality increased by 30.7% and female mortality rose by 23.8%, which actually caused the gender mortality gap to shrink. The relative symmetry in SHF mortality trends contrasts with the late divergence seen in DHF mortality, suggesting different underlying factors may be driving outcomes in each phenotype.

Finally, this study reveals a notable increase in age-adjusted mortality rates for both systolic heart failure and diastolic heart failure across certain racial groups from 2016 to 2020. While SHF consistently exhibited higher mortality than DHF across the study period, the gap narrowed significantly in several populations, particularly among Black, Hispanic, Native American, and Other racial groups. Importantly, the contribution analysis demonstrated that most of the cumulative mortality increases from 2016 to 2020 occurred during the final year, emphasizing the outsized impact of the 2020 data point. The timing strongly implicates that systemic disruptions during the COVID-19 pandemic were contributing, and the fact that minority populations (Hispanic, Native, and Black individuals) exhibited the steepest mortality surges highlights persistent healthcare inequities and the compounding effects of structural barriers during public health crises. The COVID-19 pandemic likely strained health systems, resulting in reduced access to outpatient care, delays in heart failure diagnosis and optimization, and interruptions in chronic disease management. These effects may have been more severe for minority populations already facing structural inequities in healthcare access, socioeconomic stressors, and a higher prevalence of comorbidities such as hypertension, diabetes, and obesity, all of which are risk factors for adverse heart failure outcomes.

Additionally, diastolic heart failure, which is often under-recognized, may have gone undermanaged during the pandemic due to its diagnostic complexity and reliance on follow-up care. The sudden spike in DHF mortality in 2020 across most racial groups, but especially in Hispanics and Native Americans, could reflect a vulnerability in managing this subtype under conditions of disrupted care. These findings underscore the importance of resilient and equitable healthcare infrastructure that can maintain continuity of care even during systemic crises. They also highlight the need for more targeted public health strategies aimed at identifying and mitigating heart failure mortality risks in under-resourced populations, especially during periods of societal disruption.

## Limitations

This study relied on administrative data from the National Inpatient Sample and ICD-10 diagnostic codes, which may be subject to miscoding and cannot capture the full clinical context, such as echocardiographic parameters or outpatient care. The analysis is also retrospective and observational, precluding causal inference about the drivers of mortality trends, including the impact of the COVID-19 pandemic. Third, the dataset is limited to hospitalized patients and may not reflect trends in community-managed heart failure. Despite these limitations, the use of a large, nationally representative sample strengthens the generalizability of our findings.

## Data Availability

NIS database is publicly available

## Acknowledgment

None

## Ethical Approval

The NIS database is publicly available without any patient identifier exempt from IRB

## References

[1] Bozkurt B, Ahmad T, Alexander KM, Baker WL, Bosak K, Breathett K, Fonarow GC, Heidenreich P, Ho JE, Hsich E, Ibrahim NE, Jones LM, Khan SS, Khazanie P, Koelling T, Krumholz HM, Khush KK, Lee C, Morris AA, Page RL, Pandey A, Piano MR, Stehlik J, Stevenson LW, Teerlink JR, Vaduganathan M and Ziaeian B. Heart Failure Epidemiology and Outcomes Statistics: A Report of the Heart Failure Society of America. Journal of Cardiac Failure 2023; 29: 1412–1451.

[2] Savarese G, Becher PM, Lund LH, Seferovic P, Rosano GMC and Coats AJS. Global burden of heart failure: a comprehensive and updated review of epidemiology. Cardiovascular Research 2023; 118: 3272–3287.

[3] Khan MS, Shahid I, Bennis A, Rakisheva A, Metra M and Butler J. Global epidemiology of heart failure. Nature Reviews Cardiology 2024; 21: 717–734.

[4] Heidenreich PA, Bozkurt B, Aguilar D, Allen LA, Byun JJ, Colvin MM, Deswal A, Drazner MH, Dunlay SM, Evers LR, Fang JC, Fedson SE, Fonarow GC, Hayek SS, Hernandez AF, Khazanie P, Kittleson MM, Lee CS, Link MS, Milano CA, Nnacheta LC, Sandhu AT, Stevenson LW, Vardeny O, Vest AR and Yancy CW. 2022 AHA/ACC/HFSA Guideline for the Management of Heart Failure: A Report of the American College of Cardiology/American Heart Association Joint Committee on Clinical Practice Guidelines. Circulation 2022; 145:

[5] Murphy SP, Ibrahim NE and Januzzi JL. Heart Failure With Reduced Ejection Fraction. JAMA 2020; 324: 488.

[6] Abraham WT, Psotka MA, Fiuzat M, Filippatos G, Lindenfeld J, Mehran R, Ambardekar AV, Carson PE, Jacob R, Januzzi JL, Konstam MA, Krucoff MW, Lewis EF, Piccini JP, Solomon SD, Stockbridge N, Teerlink JR, Unger EF, Zeitler EP, Anker SD and O’Connor CM. Standardized definitions for evaluation of heart failure therapies: scientific expert panel from the Heart Failure Collaboratory and Academic Research Consortium. European Journal of Heart Failure 2020; 22: 2175–2186.

[7] Kozman K, Ferrannini G, Benson L, Dahlström U, Hage C, Savarese G, Shahim B and Lund LH. Etiology of Heart Failure Across the Ejection Fraction Spectrum and Association With Prognosis. JACC: Heart Failure 2025; 13: 102491.

[8] Katz AM and Rolett EL. Heart failure: when form fails to follow function. European Heart Journal 2016; 37: 449–454.

[9] Triposkiadis F, Briasoulis A, Kitai T, Magouliotis D, Athanasiou T, Skoularigis J and Xanthopoulos A. The sympathetic nervous system in heart failure revisited. Heart Failure Reviews 2023; 29: 355–365.

[10] Bloom MW, Greenberg B, Jaarsma T, Januzzi JL, Lam CSP, Maggioni AP, Trochu J-N and Butler J. Heart failure with reduced ejection fraction. Nature Reviews Disease Primers 2017; 3: 17058.

[11] Redfield MM and Borlaug BA. Heart Failure With Preserved Ejection Fraction. JAMA 2023; 329: 827.

[12] Peikert A, Fontana M, Solomon SD and Thum T. Left ventricular hypertrophy and myocardial fibrosis in heart failure with preserved ejection fraction: mechanisms and treatment. European Heart Journal 2025;

[13] Cannata A and McDonagh TA. Heart Failure with Preserved Ejection Fraction. New England Journal of Medicine 2025; 392: 173–184.

[14] Avula HR, Leong TK, Lee KK, Sung SH and Go AS. Long-Term Outcomes of Adults With Heart Failure by Left Ventricular Systolic Function Status. The American Journal of Cardiology 2018; 122: 1008–1016.

[15] Kondo T, Henderson AD, Docherty KF, Jhund PS, Vaduganathan M, Solomon SD and McMurray JJV. Why Have We Not Been Able to Demonstrate Reduced Mortality in Patients With HFmrEF/HFpEF? Journal of the American College of Cardiology 2024; 84: 2233–2240.

[16] Vohra AS, Moghtaderi A, Luo Q, Magid DJ, Black B, Masoudi FA and Kini V. Trends in Mortality After Incident Hospitalization for Heart Failure Among Medicare Beneficiaries. JAMA Network Open 2024; 7: e2428964.

[17] Lam CSP, Arnott C, Beale AL, Chandramouli C, Hilfiker-Kleiner D, Kaye DM, Ky B, Santema BT, Sliwa K and Voors AA. Sex differences in heart failure. European Heart Journal 2019; 40: 3859–3868c.

[18] Mansur ADP, Del Carlo CH, Gonçalinho GHF, Avakian SD, Ribeiro LC, Ianni BM, Fernandes F, César LAM, Bocchi EA and Pereira-Barretto AC. Sex Differences in Heart Failure Mortality with Preserved, Mildly Reduced and Reduced Ejection Fraction: A Retrospective, Single-Center, Large-Cohort Study. International Journal of Environmental Research and Public Health 2022; 19: 16171.

[19] Shah NS, Molsberry R, Rana JS, Sidney S, Capewell S, O’Flaherty M, Carnethon M, Lloyd-Jones DM and Khan SS. Heterogeneous trends in burden of heart disease mortality by subtypes in the United States, 1999-2018: observational analysis of vital statistics. BMJ 2020; m2688.

[20] Desai RJ, Mahesri M, Chin K, Levin R, Lahoz R, Studer R, Vaduganathan M and Patorno E. Epidemiologic Characterization of Heart Failure with Reduced or Preserved Ejection Fraction Populations Identified Using Medicare Claims. The American Journal of Medicine 2021; 134: e241–e251.

[21] Alvarez-Garcia J, Lee S, Gupta A, Cagliostro M, Joshi AA, Rivas-Lasarte M, Contreras J, Mitter SS, Larocca G, Tlachi P, Brunjes D, Glicksberg BS, Levin MA, Nadkarni G, Fayad Z, Fuster V, Mancini D and Lala A. Prognostic Impact of Prior Heart Failure in Patients Hospitalized With COVID-19. Journal of the American College of Cardiology 2020; 76: 2334–2348.

